# Exploring the role of Large Language Models in Melanoma: a Systemic Review

**DOI:** 10.1101/2024.09.23.24314213

**Authors:** Mor Zarfati, Girish N Nadkarni, Benjamin S Glicksberg, Moti Harats, Shoshana Greenberger, Eyal Klang, Shelly Soffer

## Abstract

**Background:** Large language models (LLMs) are gaining recognition across various medical fields; however, their specific role in dermatology, particularly in melanoma care, is not well-defined. This systematic review evaluates the current applications, advantages, and challenges associated with the use of LLMs in melanoma care.

**Methods:** We conducted a systematic search of PubMed and Scopus databases for studies published up to July 23, 2024, focusing on the application of LLMs in melanoma. Identified studies were categorized into three subgroups: patient education, diagnosis and clinical management. The review process adhered to PRISMA guidelines, and the risk of bias was assessed using the modified QUADAS-2 tool.

**Results:** Nine studies met the inclusion criteria. Five studies compared various LLM models, while four focused on ChatGPT. Three studies specifically examined multi-modal LLMs. In the realm of patient education, ChatGPT demonstrated high accuracy, though it often surpassed the recommended readability levels for patient comprehension. In diagnosis applications, multi- modal LLMs like GPT-4V showed capabilities in distinguishing melanoma from benign lesions. However, the diagnostic accuracy varied considerably, influenced by factors such as the quality and diversity of training data, image resolution, and the models’ ability to integrate clinical context. Regarding management advice, one study found that ChatGPT provided more reliable management advice compared to other LLMs, yet all models lacked depth and specificity for individualized decision-making.

**Conclusions:** LLMs, particularly multimodal models, show potential in improving melanoma care through patient education, diagnosis, and management advice. However, current LLM applications require further refinement and validation to confirm their clinical utility. Future studies should explore fine-tuning these models on large dermatological databases and incorporate expert knowledge.

## INTRODUCTION

Large language models (LLMs), including ChatGPT, Gemini and Llama, are artificial intelligence (AI) models designed to understand and generate human-like text.^1^ These models are gaining recognition across various medical specialties for their potential to assist with clinical tasks.^2–7^ However, their specific role in dermatology, particularly in melanoma care, remains under investigation. ^8^ Multi-modal LLMs, such as GPT-4 Vision (GPT-4V), further expand this potential by combining visual and textual data. This capability could improve applications in medical imaging and diagnosis.^9^

Previous studies have shown mixed results, leading to caution among dermatologists regarding the use of these models.^10^ Nevertheless, with appropriate optimization, LLMs may improve melanoma diagnosis, patient communication, and treatment outcomes.

This systematic review aims to evaluate the current applications, advantages, and challenges associated with the use of LLMs in melanoma care.

## FOUNDATIONAL CONCEPTS

Below are the key concepts related to LLMs and their applications in healthcare. In **Figure 1**, we present a hierarchy diagram of AI terms.

**Figure 1.**
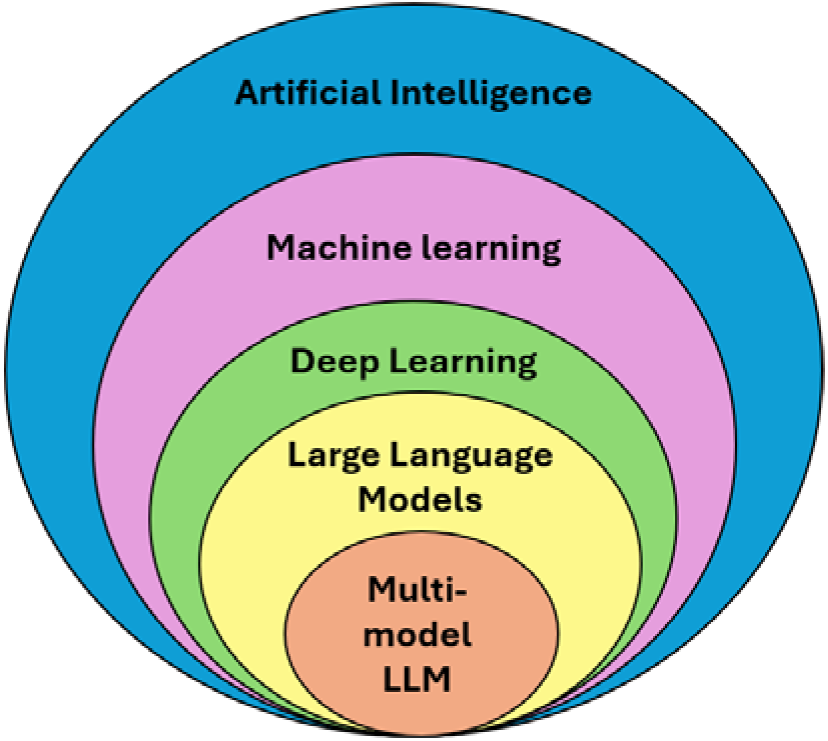
Hierarchy diagram of artificial intelligence (AI) terms.

### Artificial Intelligence and Deep Learning

AI refers to the development of algorithms capable of performing tasks that typically require human intelligence. Examples include language comprehension and image pattern recognition. Deep learning is a subset of AI that employs artificial neural networks to analyse different types of data and learn from it.^11,12^

### Artificial Neural Networks

Artificial neural networks form the foundation of deep learning. Inspired by biological neural networks, they consist of interconnected nodes, or “neurons,” organized in layers. Each neuron receives inputs, processes them, and passes an output to the subsequent layer. Each neuron is a simple computational unit, similar to a single logistic regression function. By adjusting the connections between neurons based on the input data, neural networks can learn to recognize patterns and generate predictions.^11^

### Large Language Models

LLMs are large deep learning models that process and generate human-like text. Composed of multiple transformer layers, these models employ an attention mechanism to selectively focus on different parts of the input data. This structure allows them to excel in tasks such as text recognition, language translation, and content generation.^13^ Notable examples of LLMs include ChatGPT by OpenAI and LLaMA by Meta.

### Multimodal Large Language Models

Multimodal LLMs extend the capabilities of traditional LLMs by incorporating multiple data modalities, such as text and images. These advanced models can analyse and interpret both visual and textual information, making them particularly valuable in fields like radiology and dermatology, where accurate diagnosis often requires the synthesis of information from diverse sources.^14^ In **Figure 2**, we present a diagram of possible uses of multimodal LLMs in dermatology.

**Figure 2.**
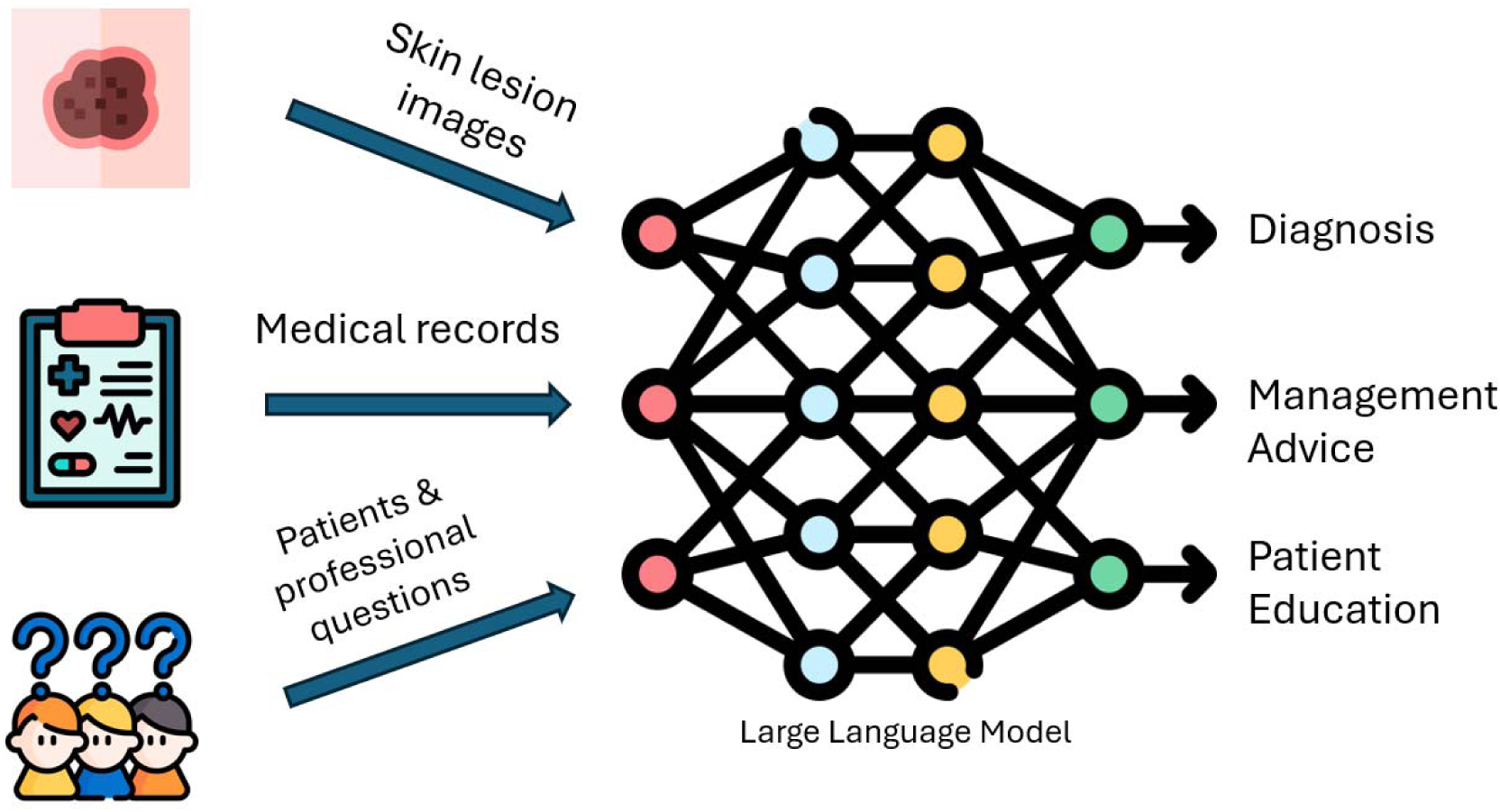
Applications of multi-modal LLMs in dermatology

## METHODS

### Search Strategy

A systematic review was conducted according to the Preferred Reporting Items for Systematic Reviews and Meta-Analyses statement (PRISMA) guidelines and the recommendations for systematic reviews of prediction models (CHARMS checklist). ^15,16^ The study is registered with PROSPERO (CRD42024575859).^17^

We searched the literature for applications of LLMs in melanoma using PubMed and Scopus. A systematic search of the published literature was conducted on July 23, 2024. Our search query was “((“Melanoma”) AND ((“ChatGPT”) OR (“large language models”) OR (“OpenAI”) OR (“Microsoft Bing”) OR (“google bard”) OR (“google gemini”)))”. To ensure thoroughness, we also reviewed the reference lists of relevant articles, but this did not yield any additional studies that met the inclusion criteria.

We excluded articles that did not specifically evaluate the application of LLMs in melanoma, non-original articles, and conference abstracts.

Study Selection:

The titles and abstracts of the identified studies were screened to determine their eligibility based on the inclusion and exclusion criteria. Any uncertainty was resolved through discussion between two reviewers, with a third reviewer consulted when necessary. The full texts of the selected articles were then independently assessed by two reviewers (MZ, SS). Discrepancies were resolved through consensus or consultation with a third reviewer (EK).

### Data Extraction

Data extraction was conducted using a standardized form to ensure consistency. Key information extracted included the first author’s name, year of publication, sample size, LLM model types, objectives, and main findings.

To investigate the specific applications and effectiveness of LLMs in different aspects of melanoma care, we divided the articles into three subgroups: patient education, clinical management, and diagnosis.

### Quality Assessment and Risk of Bias

To evaluate the risk of bias, we used the adapted version of the Quality Assessment of Diagnostic Accuracy Studies criteria (QUADAS-2).^18^

## RESULTS

Our literature search yielded a total of 45 articles from PubMed and Scopus. After the removal of 9 duplicates, the screening process found 9 studies that met our inclusion criteria. We did not identify additional studies via reference screening.^19–27^ The process of study selection and the screening methodology are detailed in the PRIZMA flow chart **(Figure 3).**

**Figure 3.**
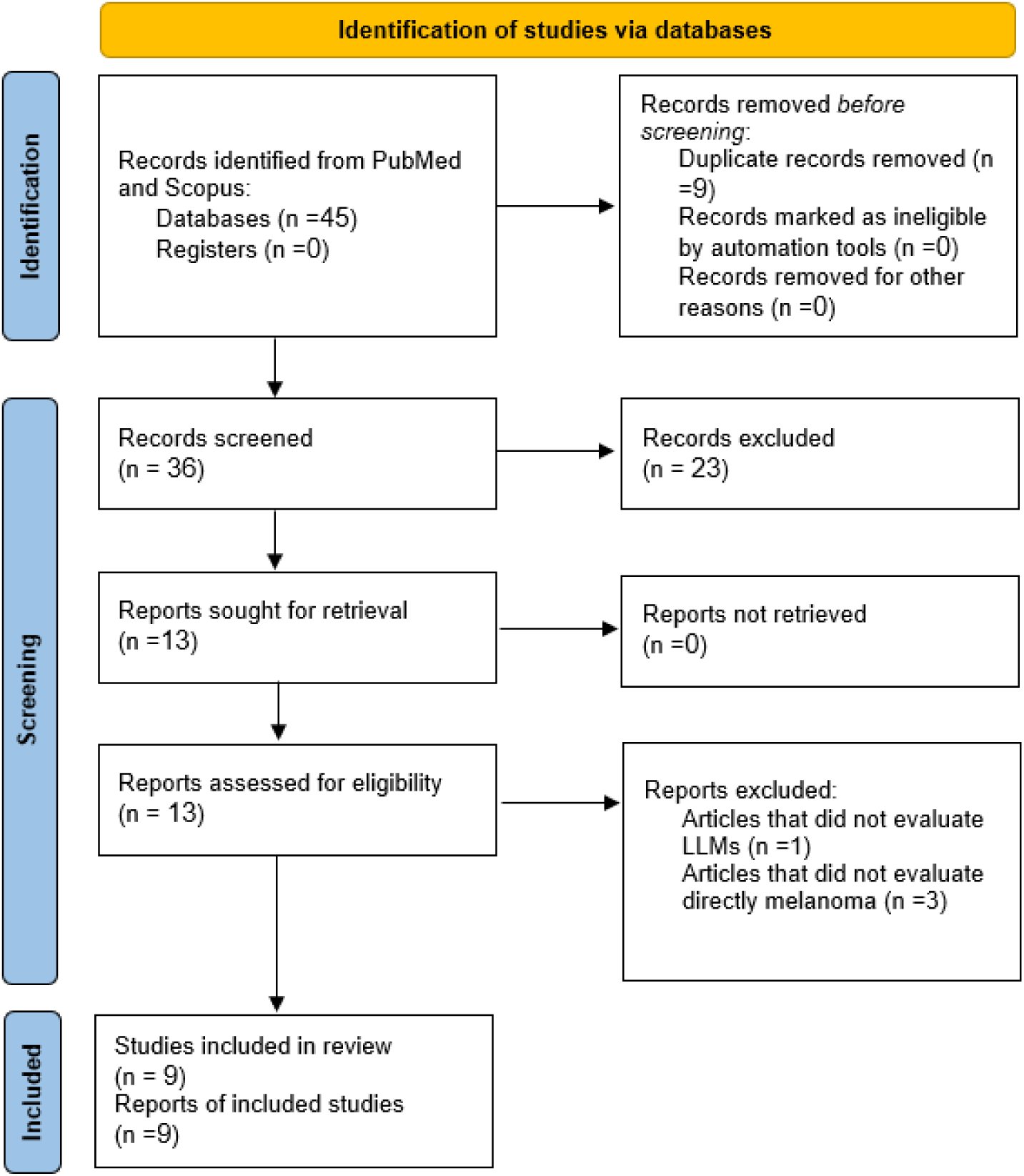
Flow Diagram of the Inclusion Process. Flow diagram of the search and inclusion process based on the Preferred Reporting Items for Systematic Reviews and Meta-Analyses (PRISMA) guidelines.

According to the QUADAS-2 tool, most papers scored as having a low to moderate risk of bias for the interpretation of the index test. A detailed assessment of the risk of bias is provided in **Supplementary Table 1.**

The characteristics of the studies are presented in **Table 1**. A summary of the objective, sample size, reference standard, main findings and conclusions are presented in **Table 2**. The main advantages and challenges in the included studies are presented in **Table 3**.

**Table 1.**
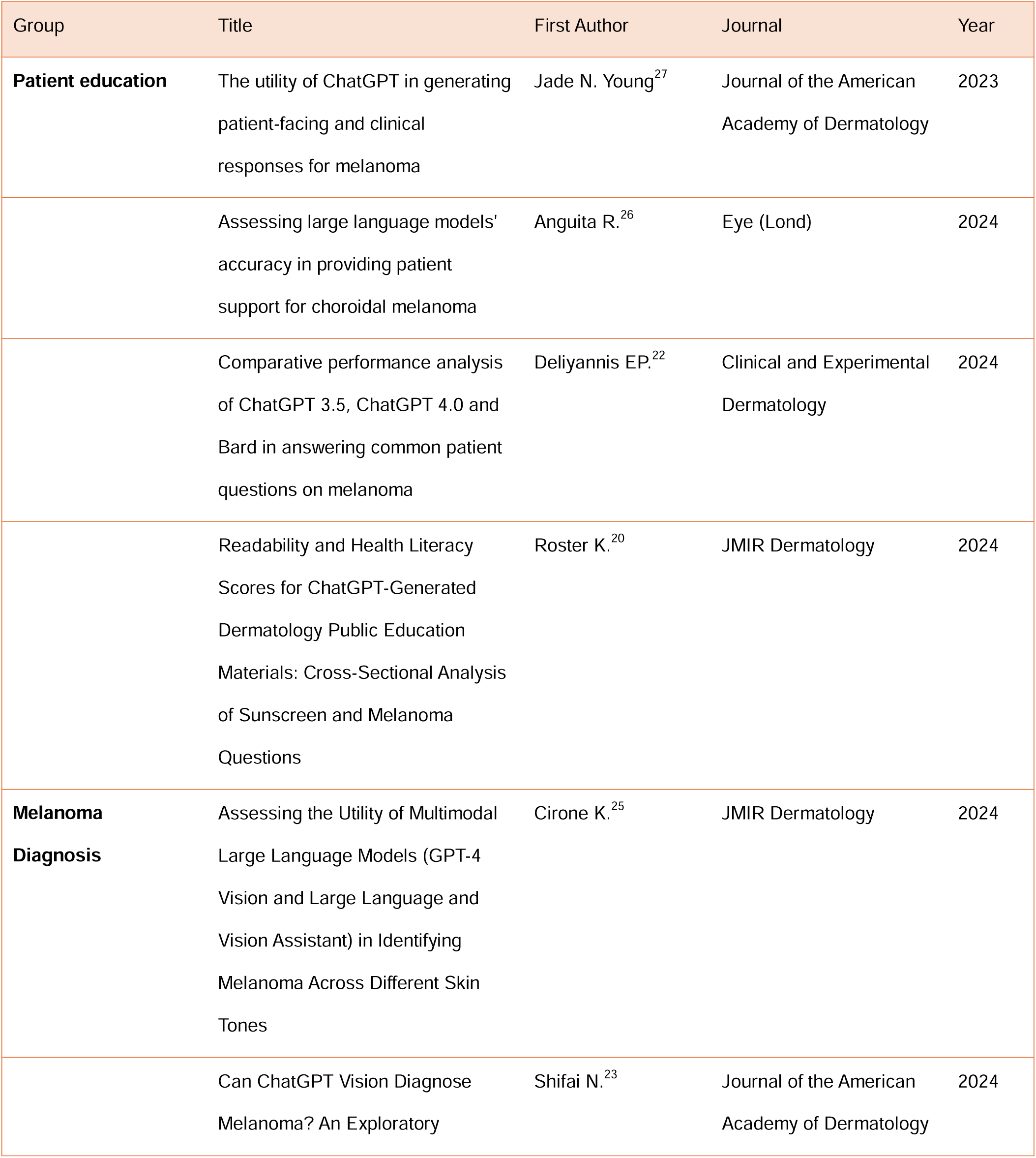
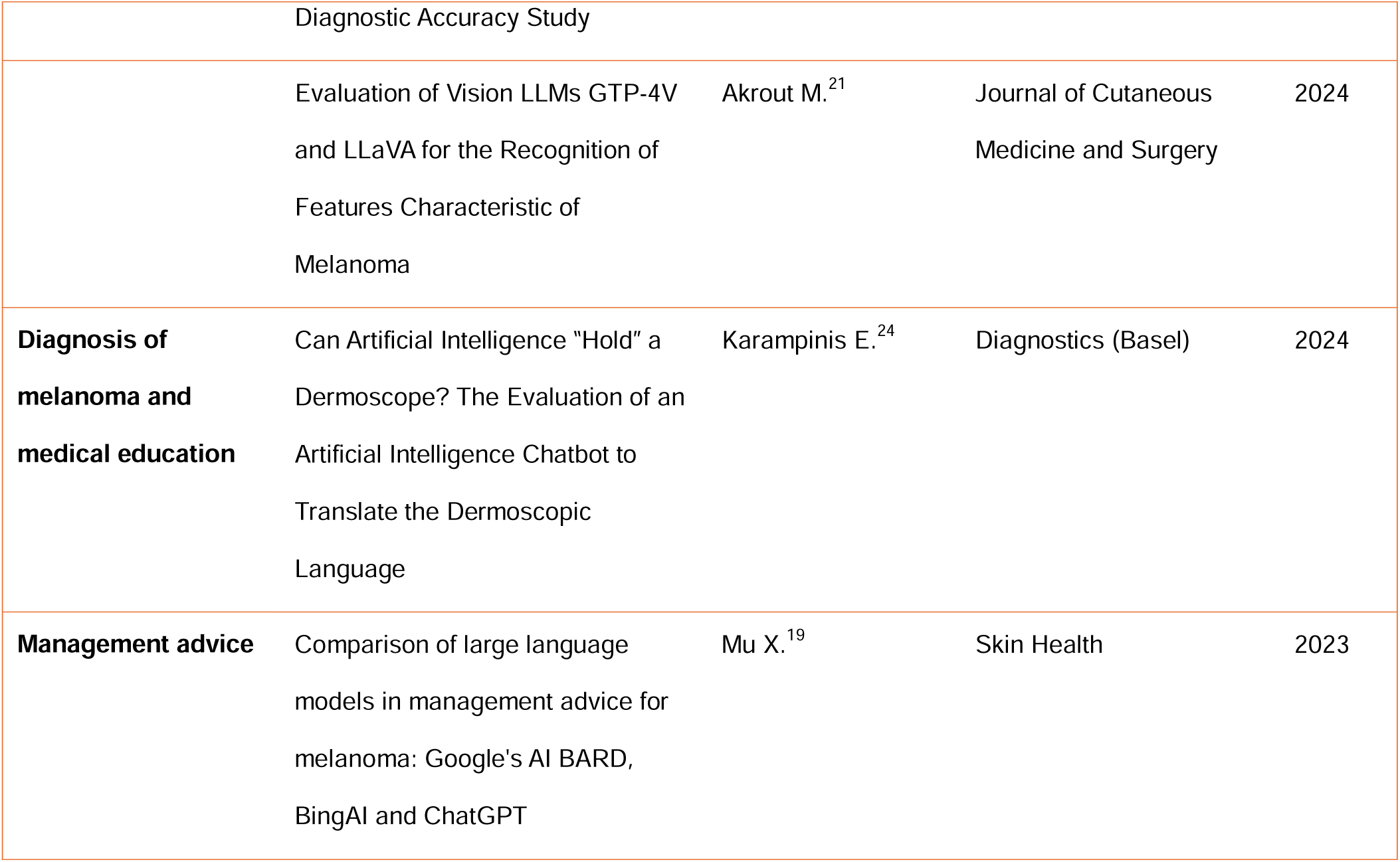
Details about reviewed articles.

**Table 2.**
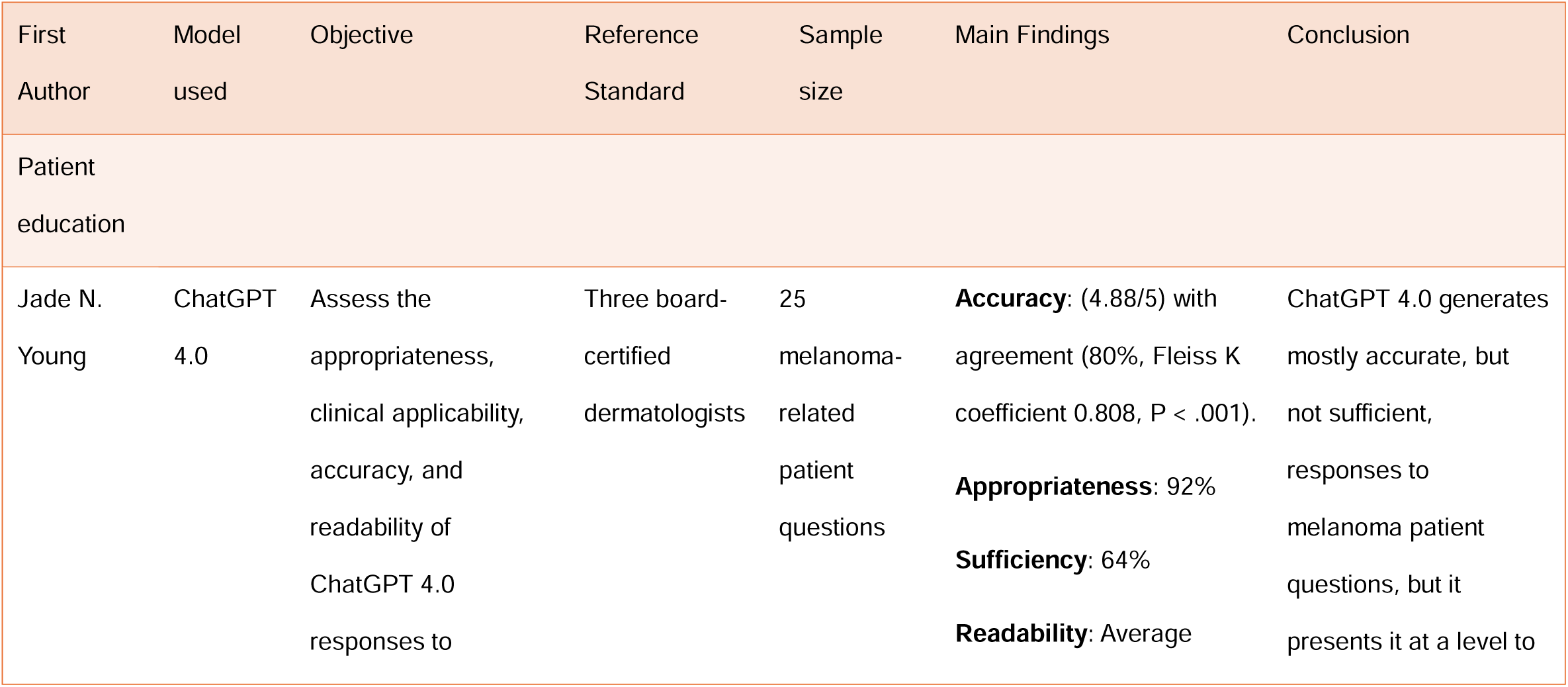
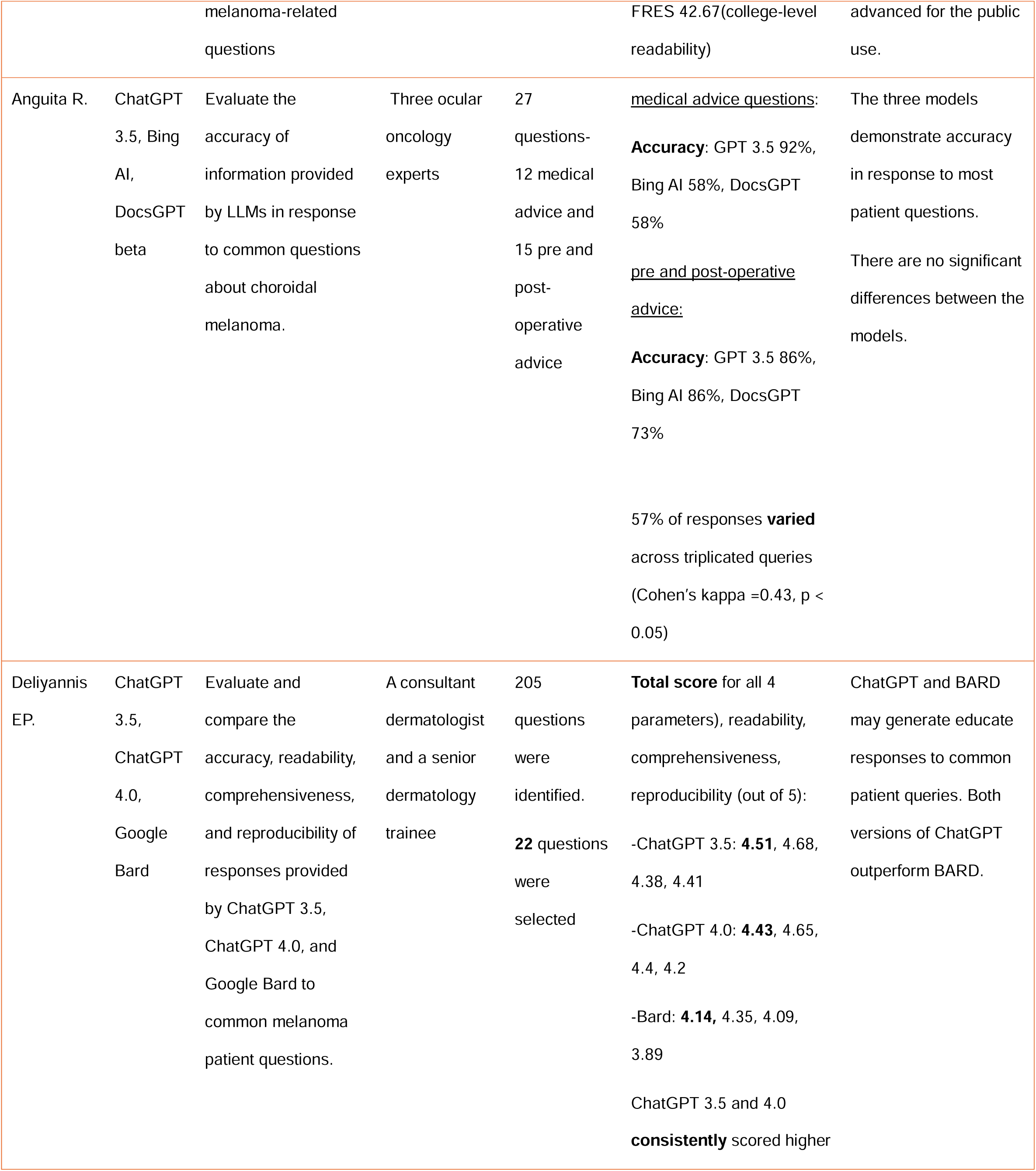
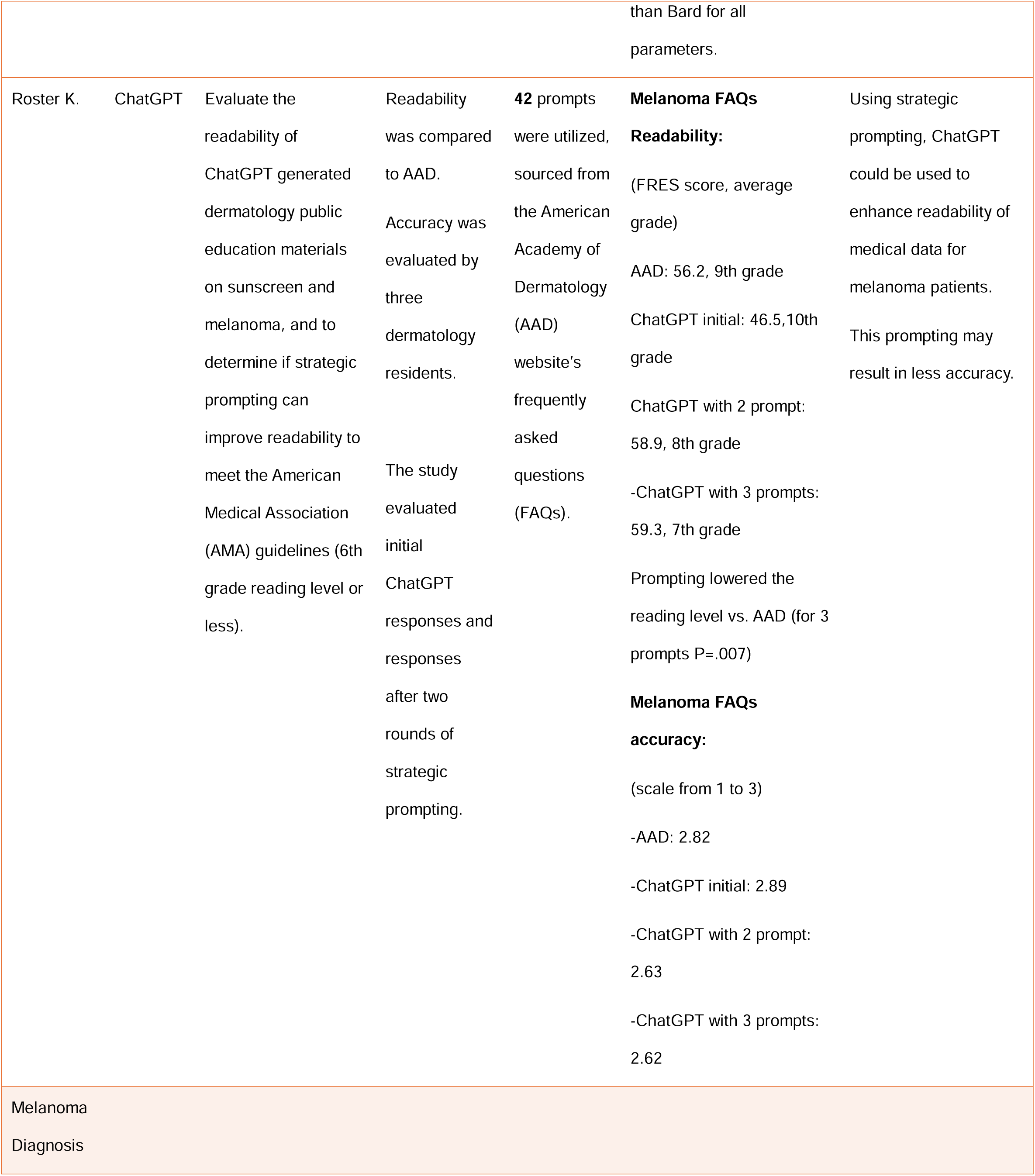
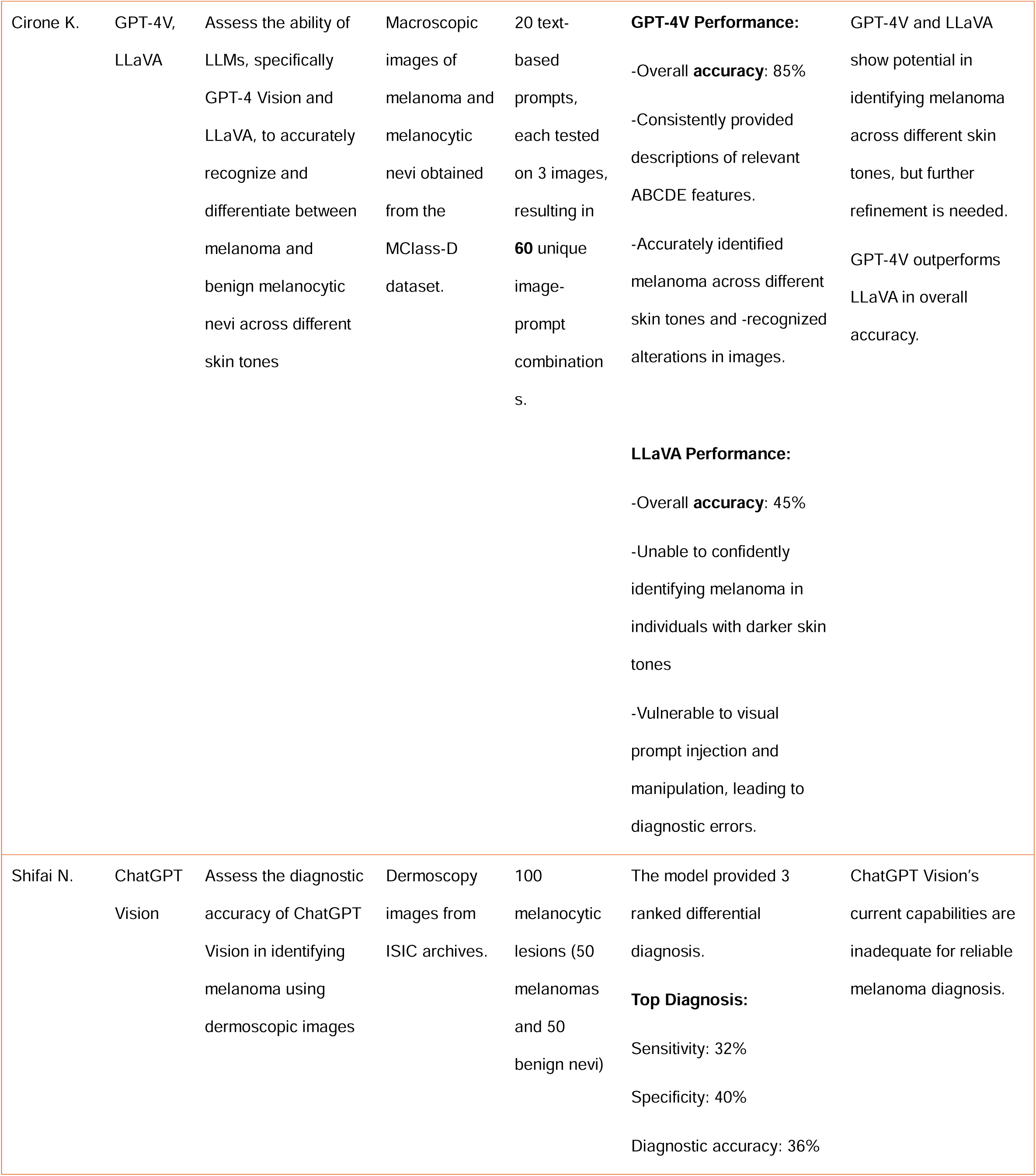
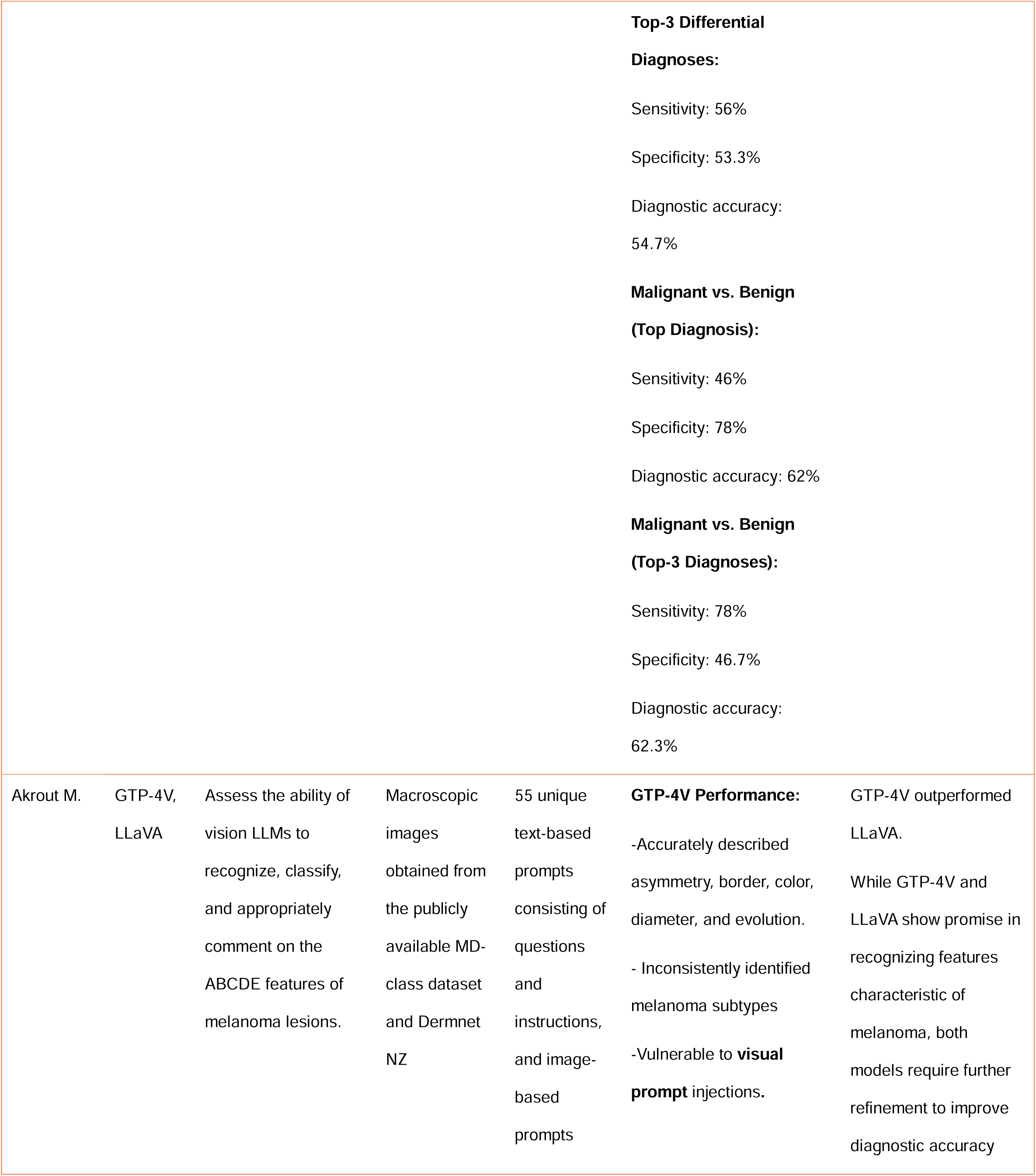
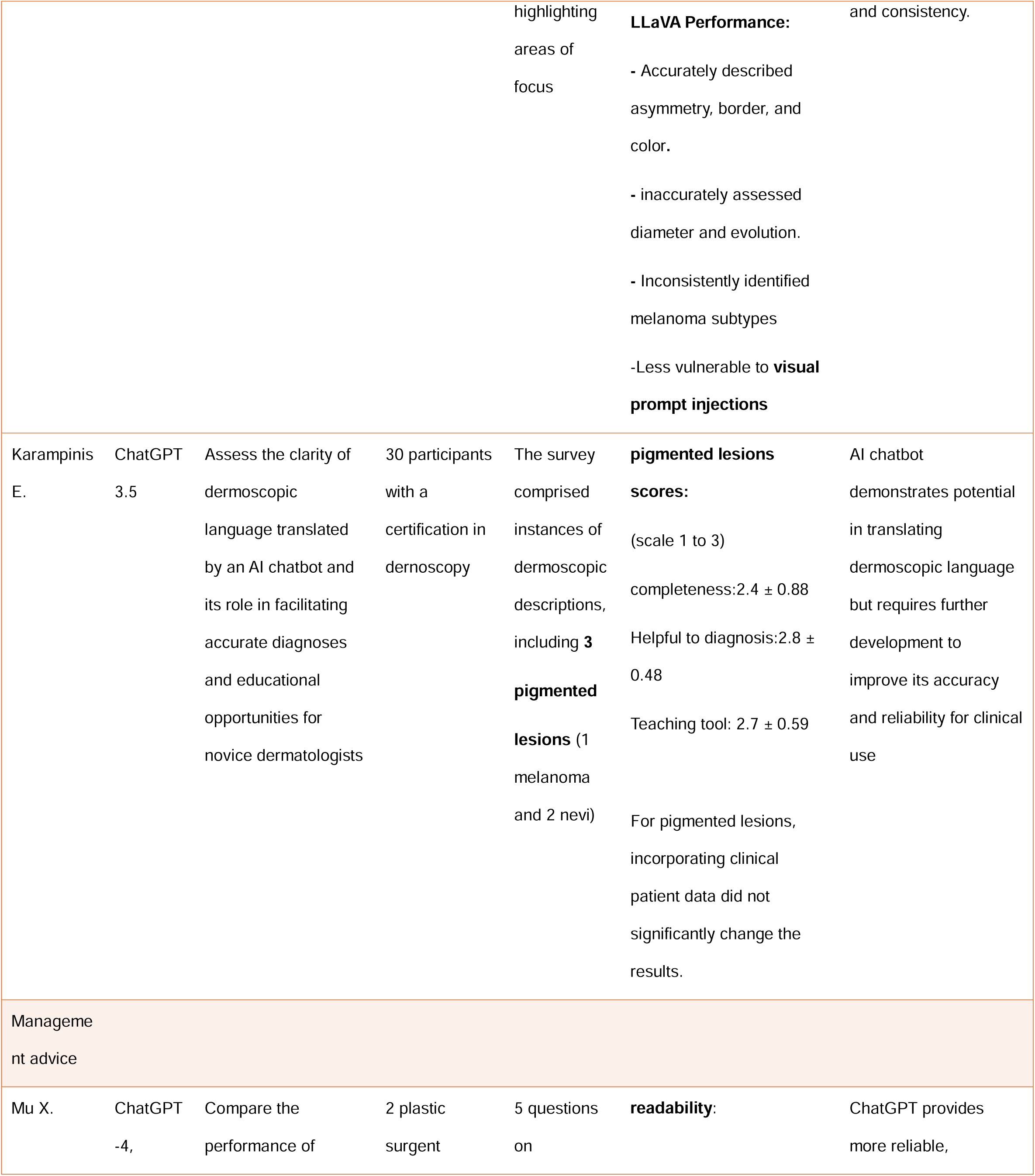
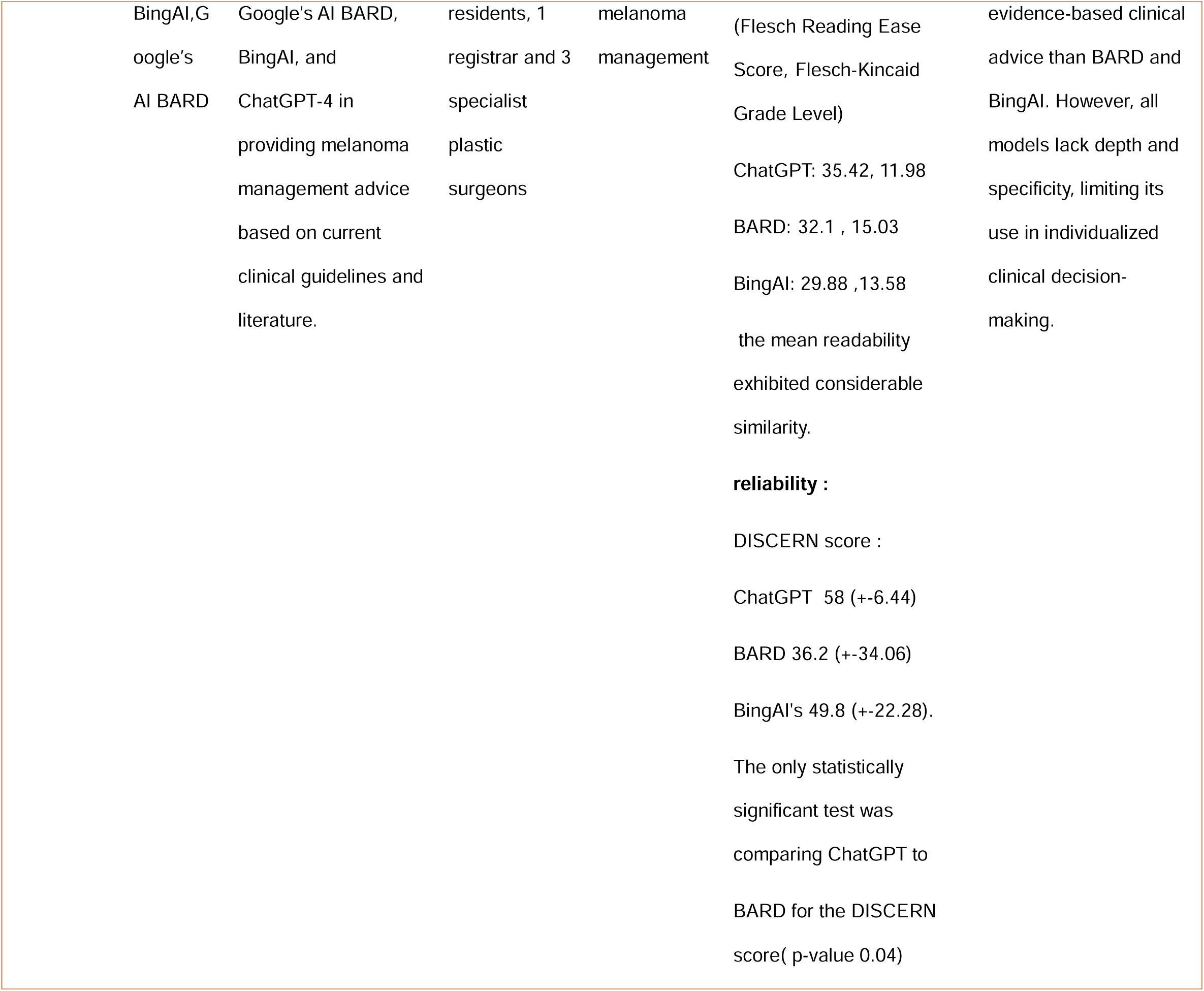
A Summary of the reviewed articles.

**Table 3:**
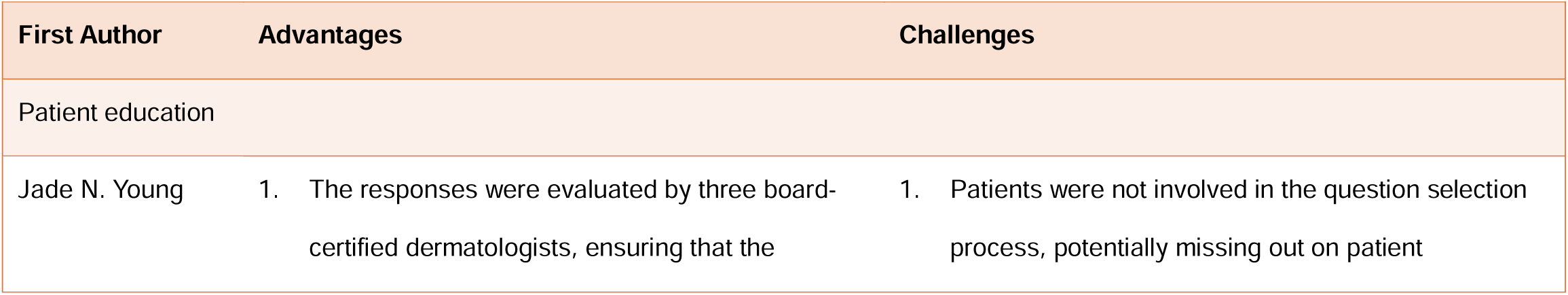
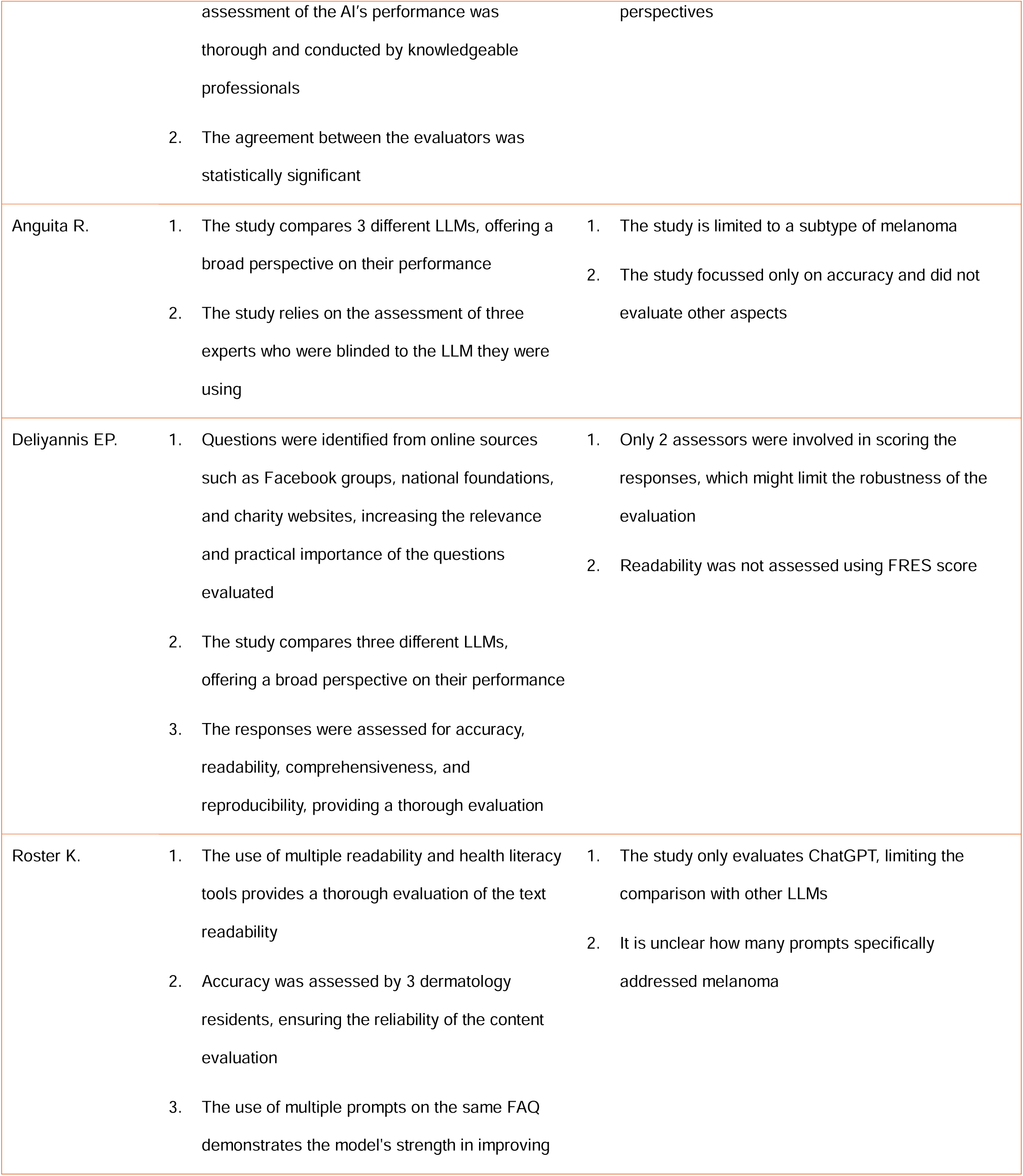
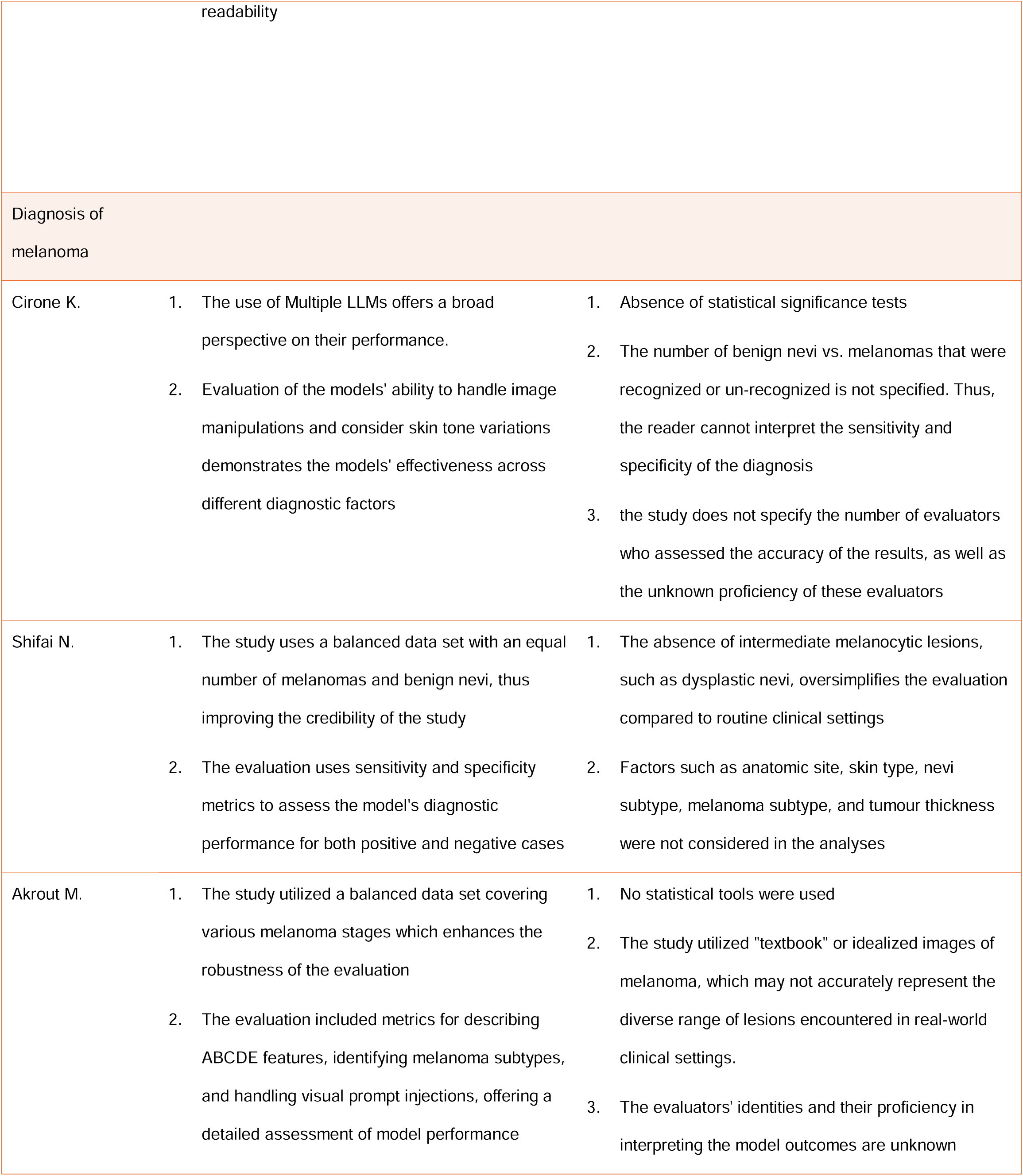
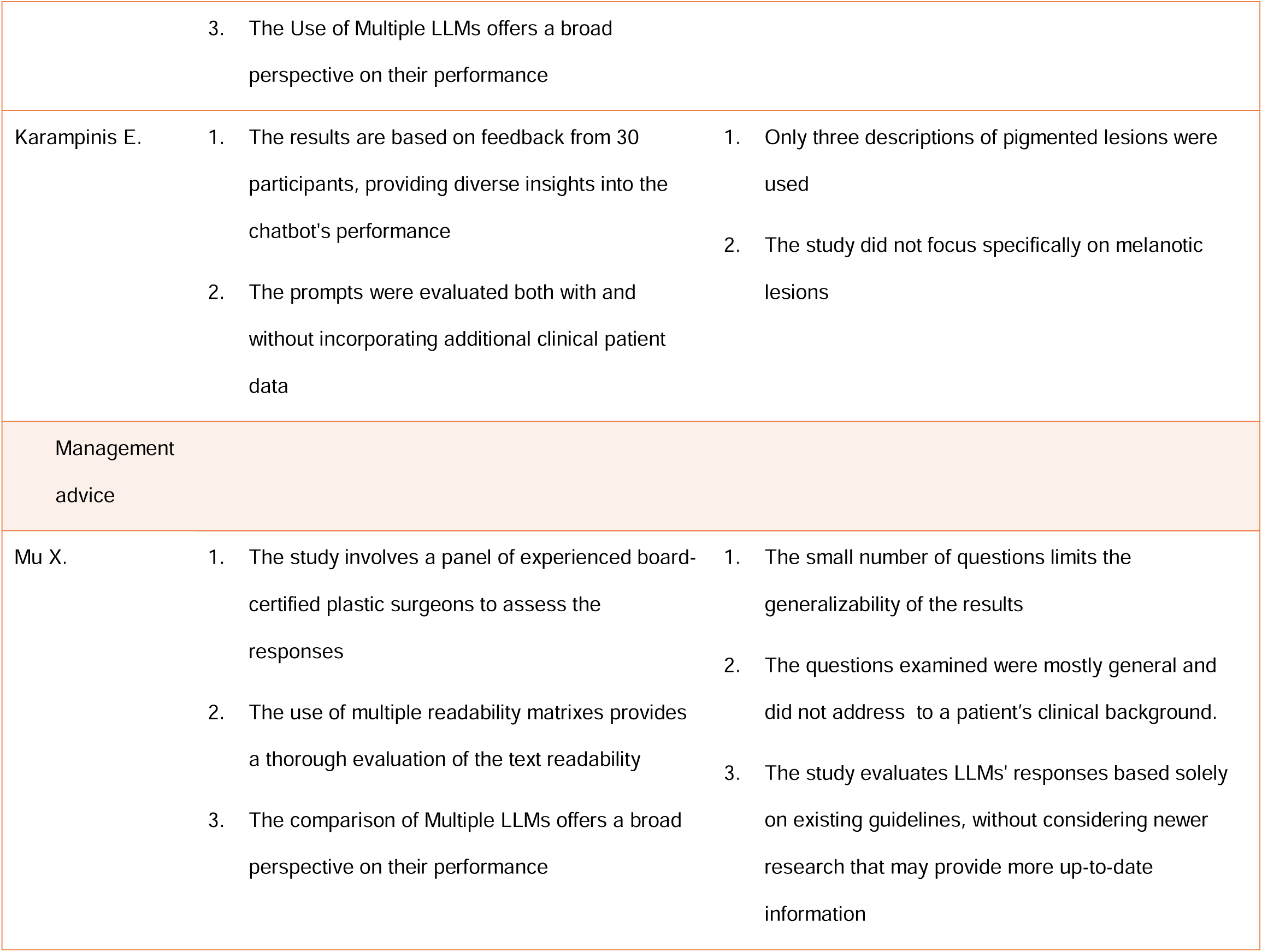
Advantages and Challenges of the reviewed articles.

Of the nine studies, five were comparative, evaluating and comparing various LLM models, such as ChatGPT, BARD, and BingAI.^19,21,22,25,26^ The remaining four studies focused on a single LLM, specifically different versions of ChatGPT.^20,23,24,27^ Three studies specifically examined multimodal LLMs, such as GPT-4V and LLaVA, highlighting their unique capabilities and associated challenges.^21,23,25^

The included studies were diverse in their objectives, methodologies, and evaluation metrics. The studies focused on the application of LLMs in melanoma diagnosis, patient education, and clinical decision-making.

### Patient education

Four studies evaluated the use of LLMs in patient education, focusing on the accuracy of responses to common patient questions.^20,22,26,27^ ChatGPT 4.0 and ChatGPT 3.5 were noted for their relatively high accuracy.

Deliyannis et al. found that while both ChatGPT and BARD can generate accurate educational responses, both ChatGPT 4.0 and 3.5 outperformed BARD.^22^ Anguita et al. focused on choroidal melanoma and found no significant accuracy differences between ChatGPT 3.5, Bing AI, and DocsGPT beta.^26^

Young et al. reported that ChatGPT 4.0 generates mostly accurate responses, scoring 4.9/5. However, only 64% of these responses were considered suitable for patient use, indicating that ChatGPT may be more effective as a supplemental tool in clinical practice. The study also found that the average readability score corresponded to a college-level comprehension, suggesting that the content might be too advanced for public use.^27^

Roster et al. addressed this readability issue by evaluating ChatGPT’s responses to questions about sunscreen and melanoma from the American Academy of Dermatology’s (AAD) website. They investigated whether prompt engineering techniques (strategic prompting) could improve readability. The study compared ChatGPT’s responses after two rounds of strategic prompting with the original answers from the AAD website. The findings showed that the initial prompt did not lower the reading level compared to the AAD content. However, with additional prompting, the reading level was reduced to 7th grade, compared to the AAD’s 9th grade level. This suggests that with proper prompt engineering, LLMs could improve the readability of medical information for melanoma patients.^20^

### Melanoma Diagnosis

Four studies examined the use of LLMs in melanoma diagnosis, focusing on their ability to identify and classify melanoma using clinical and dermoscopic data.^21,23–25^ Multi-modal LLMs, such as GPT-4V and LLaVA, played a key role in the majority of these evaluations.

Cirone et al. assessed GPT-4V and LLaVA, emphasizing their ability to integrate visual and textual data. GPT-4V demonstrated superior performance, with an overall accuracy of 85%, compared to 45% for LLaVA. Notably, LLaVA had difficulty recognizing melanoma in skin of color, unlike GPT-4V.^25^ This finding is consistent with those of Akrout et al., who also showed that GPT-4V outperformed LLaVA across all assessed features, though both models require further refinement to enhance diagnostic accuracy.^21^

This suggests that ChatGPT Vision may not yet be suitable for independent clinical use without additional refinement.

### Management advice

Only one study specifically evaluated the use of LLMs in providing melanoma management advice. Mu et al. conducted a comparative analysis of several LLMs (ChatGPT 4.0, BARD and BingAI) to assess their performance in this context. The study used five prompts related to melanoma management. ChatGPT 4.0 consistently provided more reliable, evidence-based clinical advice, outperforming the other models, with significant differences noted compared to BARD and marginally compared to BingAI. However, none of the models evaluated the risks and benefits associated with their recommendations. The limited number of questions restricts the generalizability of the findings. ^19^

## DISCUSSION

This review’s findings underscore the potential of LLMs across various domains in melanoma care, including patient education, disease diagnosis and management advice. Of particular interest is the emergence of multi-modal LLMs, which integrate visual and textual data to address the complexities of medical imaging and clinical decision-making.

In patient education, LLMs demonstrated ability to generate accurate and readable responses to common melanoma-related queries. For example, Roster et al. showed that strategic prompting can enhance the readability of ChatGPT’s outputs.^20^ This finding suggests that with appropriate fine-tuning, LLMs could become valuable tools for creating accessible patient education materials, enabling individuals to make informed decisions.

In melanoma diagnosis, multi-modal LLMs such as GPT-4V and LLaVA exhibited capabilities in distinguishing melanoma from benign lesions. Cirone et al. and Akrout et al. demonstrated GPT- 4V’s superior performance,^21,25^ particularly in handling variations in skin tone and image manipulations.^25^ Zhou et al. presented SkinGPT-4, a multi-modal LLM trained on a large collection of skin disease images and clinical notes. SkinGPT-4 demonstrated the ability to accurately diagnose various skin conditions and provide interactive treatment recommendations.^28^ In addition to LLMs, AI-based methods, particularly those utilizing dermoscopic images, have shown promising results in assisting with melanoma detection. A systematic review by Patel et al. found that AI-based algorithms achieved higher ROC (>80%) compared to dermatologists in the detection of melanoma using dermoscopic images.^29^ However, it is important to recognize that multi-modal LLMs are not yet reliable for independent clinical use. Their performance may be influenced by factors such as dataset limitations, image quality, and the lack of clinical context.

Despite these limitations, multi-modal LLMs may hold promise for applications in medical education. Sorin et al. explored the potential of multi-modal LLMs in ophthalmology education, suggesting that they could significantly impact this field by providing detailed explanations of ocular examination and imaging findings.^30^ Similarly, in the context of melanoma and dermatology, multi-modal LLMs could assist students in identifying and describing lesion characteristics, considering differential diagnoses, and developing their clinical reasoning skills.

Mu et al. investigated the use of LLMs for management advice and found that ChatGPT provided more reliable and evidence-based recommendations compared to BARD and BingAI. However, all models were limited by a lack of depth and specificity, reducing their utility in individualized clinical decision-making.^19^ This finding emphasizes the need for further refinement and validation of LLMs to ensure their recommendations align with clinical guidelines.

The limitations of this review include the small number of studies, heterogeneity in methodologies, and variations in evaluation metrics. Additionally, most studies had small sample sizes and did not involve patients in the question selection process. Furthermore, most studies focused on general melanoma questions rather than specific clinical scenarios.

In conclusion, this review highlights the potential of LLMs, particularly multi-modal models, in improving melanoma care through patient education, diagnosis, and management advice.

Despite promising results, current LLM applications require further refinement to ensure clinical utility. Future studies should explore fine-tuning these models on large dermatological databases and incorporate expert knowledge.

## Data Availability

All data produced in the present work are contained in the manuscript

